# Dietary practices and associated factors among Debre Berhan university students in Ethiopia

**DOI:** 10.1101/2025.07.24.25332094

**Authors:** Besufekad Mulugeta Urgie, Esubalew Tesfahun, Ermiyas Endewunet Melaku, Fitsum Zekarias Mohammed

## Abstract

**Background:** The transition of young people from high school to university is associated with increased autonomy, including dietary choices. However, data on this issue is scarce in the current study area.

**Purpose:** This study aimed to identify dietary practices and associated factors among university students at Debre Berhan University, Ethiopia, 2023.

**Methods:** An institution-based cross-sectional study was carried out to select 771 students using a cluster sampling technique. Then, data were collected, loaded into Epi data version 3.1 and analyzed via SPSS version 20.0. Descriptive analyses as well as bi-variable and multivariable logistic regression analyses were performed to assess the strength of the associations between the variables. Adjusted odds ratios with 95% confidence intervals and p-values < 0.05 were used to assess the level of significance.

**Results:** In this study, the prevalence of poor dietary practice was high at 77.3% at 95% CI (74.4-80.2). After adjusting the variables using logistic regression, variables such as using university meal service [AOR = 0.07, 95% CI (0.02, 0.22)], monthly pocket money < 500 birr [AOR = 0.29, 95% CI (0.13, 0.64), university cafe as a main source of food [AOR = 0.02, 95% CI (0.01, 0.05)], and 1-2 times eating of food out of student cafeteria [AOR = 0.20, 95% CI (0.06-0.68)] were associated with poor dietary practice.

**Conclusion:** Poor dietary practice among university students was high in this study. Therefore, the government and other responsible bodies should try to increase institutional-based nutritional education, create awareness, and improve campus meals containing different food groups.

## Introduction

Poor dietary practices contribute to all forms of malnutrition and diet-related non-communicable diseases (NCDs) [1]. According to the 2019 global burden of disease, dietary risk factors such as low intakes of fruits, vegetables, legumes, and whole grains and a high intake of processed meat are among the top 5 causes of mortality [2]. The transition of young people from high school to university is associated with increased autonomy, including dietary choices, small food budgets, and exposure to new social groups and food cultures that are often characterized by a shift in the composition and quality of the diet [3, 4]. These dietary practices of university students will carry over into later life, playing a long-term role in dietary behavior and food consumption, as this is an essential period in habit formation [1, 3]. As a result, there has been a growing interest in the dietary habits of adolescents and young adults recently [6].

The dietary practices and eating habits of university students are significantly altered by several factors, including exposure to stress, which leads to increased or decreased food intake and spending more time on studies [7]. Studies have shown that among university students, there is a high prevalence of poor dietary practices and risk factors for cardiovascular and metabolic diseases [6, 8, 9]. These poor dietary practices are characterized by frequent meal skipping, snacking, low fruit, vegetable and milk consumption, and increased alcohol and sugar intake [4, 6, 10-12].

In 2022, the Ministry of Health introduced food-based dietary guidelines (FBDG) that promote a healthy diet and intend to change living styles in the wider population [13]. However, data on the dietary practices and lifestyle patterns of young people are scarce. Although students are the future of a given nation, dietary practice among university students is a neglected public health issue, and the lack of a sense of importance within the general public, little economic investment at a government level, and absence of advocacy and nutrition awareness are worrisome. Therefore, recognizing and intervening the gap in dietary practices of these young population could be a way to prevent the malnutrition and diet-related non-communicable diseases (NCDs) and reduce mortalities and morbidities in the later age.

In Ethiopia, university students’ meals are prepared in university cafeterias, and most undergraduate students eat at college dining facilities with limited healthy food options. In addition, strong national studies on dietary practices among this young population are largely missing. Therefore, this study was conducted at Debre Berhan University Hospital, northeast Ethiopia.

The outcomes of the current study can benefit various stakeholders, including policymakers, academic institutions, organizations working on nutrition, and future researchers. This information can be used as additional evidence for planning and implementing intervention strategies to prevent poor dietary practices and their consequences on the community. These findings can inspire new approaches to foster activities concerning dietary practices among university students. Furthermore, this study can provide an initial idea for planning further prospective and experimental studies to explore this topic in greater depth.

## Methods and Materials

### Study design and setting

An institution-based cross-sectional study was carried out at Debre Berhan University from April 1, 2023 to May 30, 2023. The university is situated 130 km northeast of the country’s capital, Addis Ababa, in Debre Berhan Town, north Shewa district of the Amhara region, Ethiopia. Debre Berhan University (DBU) is one of the government-owned higher education institutions in Ethiopia, which was established in May, 2004 G.C. Currently, the university has enrolled a total of 19,383 students, of whom 6,759 are regular, 2,983 are extensions, 8,462 are summer, and 1,179 continue and distance education programs, under its seven colleges. There are university-owned student cafeterias located inside the university that provide meal service. Additionally, there are student lounges, and small restaurants and cafés are available around the university compound where students pay and get food. Moreover, communities are selling different types of fast food, fried food, and cooked cereals and legumes on the street near the university. Therefore, students could get food from multiple sources and they may eat healthy and unhealthy stuff.

### Population and eligibility criteria

The source population included all Debre Berhan University regular undergraduate students. The study population was randomly selected Debre Berhan University regular undergraduate students from selected colleges. Study subjects were selected from selected departments. New (first-year students) were excluded from the study.

### Sample size

The sample size (n) required for the study was calculated using the formula below.

In this formula, N is 6,759 (total number of regular undergraduate students), P is the proportion of students with good dietary habits and students who decided to eat variety food important for health, d is marginal error = 0.05 and Z1-α/2= confidence interval (95%) = 1.96. We considered p = 0.392 (39.2%) from a previous study done in Jigjig University [21]. The sample size was calculated as a population proportion formula:

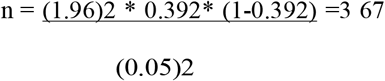

Design effect of 2 was used (due to cluster sampling): 367 * 2 = 734; non-response rate 5%, was used: 734 * 0.05 = 36.7 + 734 = 771

### Sampling procedure

A cluster sampling method was used to select 771 study participants. Four colleges were selected by simple random sampling. As each college has a different department and the sample size calculated was allocated proportionally, students were selected by a systematic random sampling method among appropriate departments using the list of students in the class. The first study participants were picked randomly at each department and every 3rd interval of the participant was selected and assigned the participant according to the number of students in the selected department.

### Study variables

The dependent variable in this study was dietary practices, and the independent variables were sociodemographic characteristics, lifestyle and behavioral practices, food group consumption and eating practices, and nutritional status (anthropometric indices).

### Operational definitions

**Snacking** refers to the intake of food between regular meals. Dietary practice Dietary practice refers to the frequency of major meals eaten in a day, where meals are eaten, snacking habits, meal skipping habits and daily intake of food types and fluid consumption [22].

**Good dietary practice** refers to eating a diversified diet that means a plate with at least four food groups in every meal and six different food groups every day out of seven food groups per day and fulfills all of a person’s nutritional needs. The six food groups in Ethiopia’s Food-Based Dietary Guidelines are as follows: 1) cereal, grains, white roots and tubers; 2) legumes; 3) nuts and oil seeds; 4) milk and dairy foods; meat, fish and egg; 5) Fruits and vegetables; 6) Fats and oils [13, 14].

**Poor dietary practice** means that a person eats fewer than five food groups per day and does not fulfill all of a person’s nutritional needs. According to Ethiopia’s food-based dietary guidelines, every person eats fewer than four food groups in every meal and six food groups every day [13, 14].

### Data measurement and the tool

The data collection was carried out using a semi-structured questionnaire. First, the questionnaire was translated from English to Amharic and back into English to ensure consistency. The final questionnaire was administered in Amharic (native language). To ensure data quality, training was provided to the data collectors (two BSc nurses) and a supervisor (one health officer). The questionnaire comprised a section to collect data on socio-demographic factors, behavioral practices, anthropometric measurement data, food group consumption and eating practices.

Before the actual data collection, the questionnaire was tested by taking 5% of the total sample size to ensure the reliability of the questioner among undergraduate students in Addis Ababa Science and Technology University (AASTU). Finally, after the data was collected, the questioners translated it back to the English language. On-spot checks, re-interviewing and checking completed questionnaires and recording quality were performed via daily supervision by the field supervisor.

### Data processing and analysis

Data were entered into Epi data version 3.1 and then exported to SPSS version 20 for further analysis. The data were coded, cleaned and checked for completeness, outliers and missing values. Descriptive analyses were conducted for the main dependent variables and all covariables considered in the study. Then, bi-variable logistic regression analysis was carried out to identify candidate variables for multi-variable logistic regression analysis, where variables with P value ≤ 0.2 were entered into the multivariable logistic regression analysis. Together, the multicollinearity test was performed using the variance inflation factor (VIF = 1.31) to check the absence of linearity. Moreover, before using the model for further interpretation, the model adequacy was checked using the Hosmer-Lemeshow (value = 0.62) goodness of the fit statistical method. Finally, variables with a P-value ≤ 0.05 in the multivariate logistic regression model were taken as statistically significant and the adjusted odds ratio along with its 95% confidence interval was considered to determine the association.

## Results

### Sociodemographic characteristics of the participants

A total of 771 students participated in this study, and the response rate was 100%. There was a nearly equal representation of 390 (50.6%) males and 381 (49.4%) females. The age of the students ranged from 19 to 36 years with a mean age of 22.2 ± 1.58 years. When we see the place of origin, 428 (55.5%) of the students were from urban areas, while 343 (44.5%) students came from rural areas. The monthly stipend of the students was < 500 ETB in 265 (34.4%) of students and 506 (65.5%) of students. The majority of the students (679, 88.1%) feed their meal from the university’s student cafeteria. When we look at the usual source of meal, the majority of the students (599; 77.7%) permanently feed from the university’s student cafeteria, and only 90 (11.7%) non-student cafeteria and 82 (10.6%) eat from both university student’s café and other sources (**Table** 1).

### Lifestyle characteristics, health behaviors, and anthropometric measurements of the study participants

From the total 771 study participants, only 296 (38.4%) had regular physical exercise. Regarding other health behaviors, 88 (11.4%) students drank alcohol, 9 (1.2%) students smoked cigarettes, and 657 (85.2%) students had no habit of drinking alcohol or smoking cigarettes. The majority of the students ate readily available meals 738 (95.72%) with comparable statistics between males and females. When we observed the regularity of meals, 552 (71.6%) of students ate their meals regularly, while 219 (28.4%) did not eat their meals regularly. The majority of the students commonly miss their breakfast (454 (58.9%) followed by dinner (195 (27.3%) and then lunch in 121 (15.7%) of the students. Concerning the number of meals per day, 563 (73.02%) of the students ate at least 3 meals a day, 118 (15.3%) ate twice a day, 47 (6.1%) ate only once a day and 43 (5.6%) of the students ate four or more times a day. Additionally, only 62 (8%) of the students ate snacks daily and 553 (71.7%) rarely ate snacks. When we look at the anthropometric measurements of the study participants, the mean body mass index (BMI) was 20.38 ± 2.73Kg/m2. Based on BMI, 205 (26.59%) of the participants were underweight; overweight and obesity was 6.35% and 0.13% respectively. Overweight and obesity was more common among female students (9.19%) than among males (0.26%), but the prevalence of underweight was high among male students 109 (27.95%) than female 96 (25.2%) (**Table 2**).

### Dietary practice of the study participants

The most consumed food categories were cereal-based foods (771 (100%), legumes (759 (98.4%), and oil and fat (97.8%), followed by vegetables (313 (40.6%), fruits 172 (22.3%), milk and milk products 151 (19.6%), and meat and eggs 129 (16.7%). Accordingly, from the 771 study participants, 77.3% had “poor” dietary practice and 22.7% had “good” dietary practice (**Table** 3).

### Factors associated with poor dietary practices among Debre Berhan University students

A binary logistic regression model was used to determine the association between dietary practice and the independent variables. Accordingly, five variables (age of the student, pocket money (stipend), source of meal, parents’ education level, mode of enrollment to university’s student cafeteria and frequency of feeding) were significantly associated with poor dietary practice among university students (P <0.2). Lastly, multivariable logistic regression analysis was used to identify confounding factors. According to this analysis, only four of the above variables (mode of enrollment in the student café, pocket money < 500 ETB, 1-2 times eating food outside the student cafeteria, and place of eating in the student cafe) had a significant association at the 5% significance level. Therefore, based on multiple logistic regression analysis, study participants who used university meal service were 92.5% less likely to have a good dietary habit as compared to non-café participants [AOR = 0.07, 95% CI (0.02-0.22)]. Participants who had monthly pocket money (stipend) < 500 ETB were 70.8% less likely to have good dietary practice compared to those who had income > 500 ETB [AOR = 0.29, 95% CI (0.13-0.64)]. Based on the main source of meal, participants who used university meal service were 97.2% less likely to have good dietary practice compared to those who used both sources [AOR = 0.028, 95% CI (0.01-0.05)]. Those participants who eat 1-2 meals a day out of the university cafeteria were 79.6% less likely to have good dietary practice compared to those who ate food out of the student cafeteria at all-time [AOR= 0.20, 95% CI (0.06-0.68)] (**Table** 4).

## Discussion

This study aimed to assess the dietary practice and its determinants among undergraduate students of Debe Berhan University by including a number of variables from various categories, such as socio-demographic characteristics, lifestyle and behavioral practice, food group consumption and eating habits, and nutritional status (anthropometric indices).

According to the findings of this study, the prevalence of poor dietary practice among university students was 77.3%. This figure was much higher than those of studies conducted in Finland [15] and Kenya [16]. The higher prevalence of poor dietary practice in the current study area might be due to the current inflation that influenced the cost of food.

In this study, poor dietary practice was significantly associated with utilization of university meal service, monthly pocket money < 500 ETB, usage of university meal as a main source of meal, and less frequent eating habit of meal outside university meal service.

Despite the well-known importance of the regular consumption of vegetables, fruits, milk and milk products, and meat and eggs, in the current study, participants ate vegetables in 313 (40.6%), fruits in 172 (22.3%), milk and milk products in 151 (19.6%) and meat and eggs in 129 (16.7%). These figures are lower than those of studies conducted in the Kingdom of Saudi Arabia [9, 17], India [18], and Sudan [19]. The explanation can be low socio-economic status, high cost of a healthy diet, absence of advocacy and little economic investment by the government for implementation of good dietary practice.

Eating regular meals including breakfast is considered a healthy eating behavior, but in this study only 552 (71.6%) participants ate meals regularly. This figure is in line with the study conducted in Universiti Brunei Darussalam [20], India [18, 21]; but higher than the study conducted in Colombia [22]; Saudi Arabia [23], and Italy [24]. Dietary guidelines, including the Ethiopian Food-Based Dietary Guidelines-2022 suggest and advice eating 3-4 servings per day. In this study, 606 (78.6%) of the study participants ate at least 3-4 meals a day, which is higher than that in the study conducted at Universiti Brunei Darussalam (52.5%) [20].

From the servings, breakfast is the most commonly 454 (58.9%) missed meal and this is in line with several studies done in Universiti Brunei Darussalam; Bahrain (50%) [19]. Additionally, only 62 (8%) of the students ate snacks daily and 553 (71.7%) rarely ate snacks.

As a strength, this study is the first to assess dietary practices among university students and it also uses a large sample size. On the other hand, one the potential limitations of this study is that the university cafeteria is the main dining place on campus where most undergraduate students who use meal service eat every day whatever is served based on the university’s meal schedule.

## Conclusions

This study showed that the prevalence of poor dietary practices among Debre Berhan university students is high (78%). This poor dietary practice is significantly associated with using university meal service, low pocket money (stipend), place of eating, frequency of eating food out of student cafeteria associate factors of dietary habits. Therefore, we recommend the need for efforts to promote good dietary practices among youths. By promoting the availability of healthy food options in schools, communities, and homes, we can help adolescents make better choices and develop a taste for nourishing foods. The findings revealed widespread unhealthy eating habits and suboptimal nutritional status among a substantial proportion of female students. Policymakers should prioritize and expand interventions aimed at enhancing dietary practices among women of reproductive age, particularly those in university settings.

## Data Availability

All data produced in the present study are available upon reasonable request to the authors

## Disclosures

### Ethics and consent

Ethical approval was issued by Ethical Clearance Review Board of Asrat Woldeyes Health Science Campus, Debre Berhan University (IRB Protocol number IRB-143). In accordance with the Helsinki Declaration guidelines and regulations, written informed consent was obtained from all subjects who participated in this study. In addition, all necessary data were collected and registered based on the unique codes of the study participants and names were not taken throughout the study. Hence, all the information was kept confidential.

### Conflicts of interest

The authors have declared that they have no conflict of interest in this study.

### Data availability statement

All data produced/used in this study are available upon reasonable request from the corresponding author

### Funding

This study did not receive any funding

## Notes

### Competing Interest Statement

The authors have declared no competing interest.

### Author Declarations

Ethical approval was issued by Ethical Clearance Review Board of the Asrat Woldeyes Health Science Campus, Debre Berhan University (IRB Protocol number IRB-143). In accordance with the Helsinki Declaration guidelines and regulations, written informed consent was obtained from all subjects who participated in this study. In addition, all necessary data were collected and registered based on the unique codes of the study participants and names were not taken throughout the study. Hence, all the information was kept confidential.

## References

1. Ronto: The global nutrition transition: trends, disease… - Google Scholar [Internet]. [cited. 2023, 204:1223–49.

2. Abraham S, Noriega BR, Shin JY: College students eating habits and knowledge of nutritional requirements. J Nutr Hum Health. 2018, 2:13–7.

3. Al-Awwad NJ, Al-Sayyed HF, Zeinah ZA, Tayyem RF: Dietary and lifestyle habits among university students at different academic years. Clin Nutr ESPEN. 2021, 44:236–42.

4. Sprake EF, Russell JM, Cecil JE, Cooper RJ, Grabowski P, Pourshahidi LK, et al.: Dietary patterns of university students in the UK: a cross-sectional study. Nutr J. 2018, 17:90.

5. Oti J, Eshun G: Dietary Habits and Nutritional Status of Undergraduate Students of Winneba Campus of University of Education, Winneba, Ghana. Jou Food SciNutri. 10920202023, 6:2020–11.

6. Alshammari GM, Osman MA, Alabdulkarem KB, Alsoghair SM, Mohammed MA, Al-Harbi LN, et al.: The effect of dietary behaviors on the nutritional status and associated factors of Yemeni students in Saudi Arabia. Plos One. 2022, 17:0268659.

7. Mogeni BK, Ouma LO: Dietary patterns, behaviors, and their associated factors among university students in coastal Kenya. Powerful Food Agric. 2022, 8:2132873.

8. Mohammed ESE: Assessment of Nutritional Status and Its Related Factors among Undergraduate Students in Juba University, Sudan East Afr Sch J Med Sci. 2020, 3:223–32.

9. Bede F, Cumber SN, Nkfusai CN, Venyuy MA, Ijang YP, Wepngong EN, et al.: Dietary habits and nutritional status of medical school students: the case of three state universities in Cameroon. Pan Afr Med J [Internet. 2020, [cited:2023-6.

10. Pop LM, Iorga M, Muraru ID, Petrariu FD: Assessment of dietary habits, physical activity and lifestyle in medical university students. Sustainability. 2021, 13:3572.

11. Kim KH: A study of the dietary habits, the nutritional knowledge and the consumption patterns of convenience foods of university students in the Gwangju area. Korean J Community Nutr. 2003, 181:91.

12. Bekele TH: Ethiopian food-based dietary guidelines: development, evaluation, and adherence monitoring [Internet]. Wageningen University and Research. 202220239,

13. Bekele TH, Trijsburg L, Brouwer ID, de Vries JH, Covic N, Kennedy G, et al.: Dietary recommendations for Ethiopians based on priority diet-related diseases and causes of death in Ethiopia: An umbrella review. Adv Nutr [Internet. 202320239,

14. El Ansari W, Suominen S, Samara A: Eating habits and dietary intake: is adherence to dietary guidelines associated with importance of healthy eating among undergraduate university students in Finland? Cent Eur J Public Health. 2015, 23:306–13.

15. Waweru G: A cross-sectional analysis of dietary practices and nutrition status of female undergraduate students at Kenyatta University, Kenya. Am J Food Sci Nutr. 2020, 2:12–20.

16. Almutairi KM, Alonazi WB, Vinluan JM, Almigbal TH, Batais MA, Alodhayani AA, et al.: Health promoting lifestyle of university students in Saudi Arabia: a cross-sectional assessment. BMC Public Health. 2018;18:1093.

17. Yadav H, Naidu S, Baliga SS, Mallapur MD: Dietary pattern of college going adolescents (17-19 years) in urban area of Belagavi. Age. 2015;17:22.

18. Musaiger AO, Awadhalla MS, Al-Mannai M, AlSawad M, Asokan GV: Dietary habits and sedentary behaviors among health science university students in Bahrain. Int J Adolesc Med Health [Internet. 201712023, 1515:2015–0038.

19. Yun TC, Ahmad SR, Quee DKS: Dietary habits and lifestyle practices among university students in Universiti Brunei Darussalam. Malays J Med Sci MJMS. 2018, 25:56.

20. Kumar CA, Revannasiddaiah N, Savanthe AM, Patel PK: Dietary Habits of Undergraduate Medical Students-A Cross-Sectional Study. 201420239.

21. Espitia-Almeida F, Mora-García M, Coquel-Bru A, Orozco-Sánchez C: Dietary habits and nutritional status in students of the university corporation Rafael Núñez, Cartagena-Colombia. Nutr Food Sci. 2022, 52:403–11.

22. Abdelhafez AI, Akhter F, Alsultan AA, Jalal SM, Ali A: Dietary practices and barriers to adherence to healthy eating among King Faisal University students. Int J Environ Res Public Health. 2020;17:8945.

23. Lupi S, Bagordo F, Stefanati A, Grassi T, Piccinni L, Bergamini M et al. Assessment oflifestyle and eating habits among undergraduate students.

