## Supplemental Table 1, 2, 3, and 4 for "Dietary practices and associated factors among Debre Berhan university students in Ethiopia"

**Table 1**: Socio-demographic characteristics of undergraduate students, Debre Berhan University, Ethiopia, 2023

| **Variable** | **Category** | **Frequency (n=771)** | **Percent (%)** |
| --- | --- | --- | --- |
| Age | < 20 years | 97 | 12.6 |
|  | 21-24 years | 630 | 81.7 |
|  | > 25 years | 44 | 5.7 |
| Sex | Male | 390 | 50.6 |
|  | Female | 381 | 49.4 |
| Residence | Rural | 343 | 44.5 |
|  | Urban | 428 | 55.5 |
| Mode of meal service from university | Use university meal | 679 | 88.1 |
|  | Don’t use | 92 | 11.3 |
| Father education | No formal education | 132 | 17.1 |
|  | No formal education but literate | 258 | 33.5 |
|  | Formal education | 81 | 49.4 |
| Mother education | No formal education | 242 | 31.4 |
|  | No formal education but literate | 242 | 31.4 |
|  | Formal education | 287 | 37.2 |
| Family monthly income | < 10,000 ETB | 99 | 77.7 |
|  | 10,001-20,000 ETB | 120 | 15.6 |
|  | > 20,001 ETB | 52 | 6.7 |
| Usual source of food | University’s student cafeteria | 599 | 77.7 |
|  | Other than university’s student cafeteria | 90 | 11.7 |
|  | Student cafeteria and other source | 82 | 10.6 |
| Year of study | 2nd year | 177 | 23.0 |
|  | 3rd year | 146 | 18.9 |
|  | 4th year | 280 | 36.3 |
| Department | Non health | 680 | 88.2 |
|  | Health | 91 | 11.8 |
| Monthly pocket money (stipend) | ≤ 500 ETB | 265 | 34.4 |
|  | > 500 ETB | 506 | 65.6 |

**Table 2**: Lifestyle characteristics, health behaviors and anthropometric measurement of the study participants, Debre Berhan University, Ethiopia, 2023, (n=717)

| **Variables** |  | **Frequency (N)** | **Percent (%)** |
| --- | --- | --- | --- |
| Regular physical exercise | Yes | 296 | 38.4 |
|  | No | 474 | 61.5 |
| Type of exercise | Football | 177 | 59.8 |
|  | Aerobic | 64 | 21.6 |
|  | Gymnastic | 76 | 25.7 |
|  | Others | 73 | 24.6 |
| Substance use | Alcohol | 88 | 11.4 |
|  | Cigarette | 9 | 1.2 |
|  | Both | 17 | 2.15 |
|  | None | 656 | 85.0 |
| Eat meals regularly | Yes | 552 | 71.6 |
|  | No | 219 | 28.4 |
| Sources of meal | Ready meals | 738 | 95.7 |
|  | Own preparation | 33 | 4.2 |
| Meals served outside student cafeteria | 1-2 meals a day | 443 | 57.4 |
|  | ≥ 3 meals a day | 148 | 12.2 |
|  | None | 119 | 15.4 |
|  | All meals | 61 | 7.9 |
| Frequency of meals a day | One | 47 | 6.1 |
|  | Two | 118 | 15.3 |
|  | Three | 563 | 73.0 |
|  | ≥ Four | 43 | 5.58 |
| Meals commonly skips | Breakfast | 454 | 58.9 |
|  | Lunch | 121 | 15.7 |
|  | Dinner | 195 | 27.3 |
| Frequency of snack | Daily | 62 | 8.0 |
|  | 3-4 times a week | 90 | 11.6 |
|  | Twice a week | 66 | 8.5 |
|  | < 2 times a week | 553 | 71.7 |
| Frequency of breakfast | Daily | 257 | 33.3 |
|  | 3-4 times a week | 402 | 52.1 |
|  | Twice a week | 111 | 14.4 |
| Frequency of lunch | Daily | 636 | 82.5 |
|  | 3-4 times a week | 95 | 12.3 |
|  | Twice a week | 16 | 2.07 |
|  | <2 times a week | 24 | 3.11 |
| Frequency of dinner | Daily | 564 | 73.1 |
|  | 3-4 times a week | 96 | 12.4 |
|  | Twice a week | 28 | 3.6 |
|  | <2 times a week | 83 | 10.7 |
| Body mass index (BMI) | Underweight (< 18.5) | 205 (26.5) | 26.5 |
|  | Normal (18.5-24.9) | 514 (66.6) | 66.6 |
|  | Overweight (25-29.9) | 50 (6.35) | 6.3 |

**Table 3**: Food group consumption and dietary practice among Debre Berhan University students, Ethiopia, 2023

| **Food group consumption** |  | **Frequency (N)** | **Percent (%)** |
| --- | --- | --- | --- |
| Do you eat vegetables daily? | Yes | 313 | 40.6 |
|  | No | 458 | 59.4 |
| Do you eat fruit daily? | Yes | 172 | 22.3 |
|  | No | 597 | 77.7 |
| Do you eat cereals daily? | Yes | 771 | 100 |
| Do you eat legumes daily? | Yes | 759 | 98.4 |
|  | No | 12 | 1.6 |
| Do you eat meat, and egg daily? | Yes | 129 | 16.7 |
|  | No | 642 | 83.3 |
| Do you drink milk and milk product daily? | Yes | 151 | 19.6 |
|  | No | 618 | 80.4 |
| How often do you take soft drink daily? | 3-4 times a day | 26 | 3.4 |
|  | 1-2 times a day | 52 | 6.7 |
|  | Once a day | 13 | 16.9 |
|  | 2-3 times a week | 371 | 48.1 |
|  | Rarely | 309 | 40.1 |
| Eat energy dense foods and fast food daily? | 3-4 times a day | 36 | 4.7 |
|  | 1-2 times a day | 48 | 6.2 |
|  | Once a day | 12 | 1.6 |
|  | 2-3 times a week | 392 | 50.8 |
|  | Rarely | 277 | 35.7 |
| Dietary practice | Good | 175 | 22.7 |
|  | Poor | 596 | 77.3 |

**Table 4**: Summary of multivariate analysis of factors associated with dietary habit among students of Debre Berhan University, Ethiopia, 2023.

| **Variable** | | **Dietary habit** | | **COR (95% CI)** | **AOR (95% CI)** |
| --- | --- | --- | --- | --- | --- |
|  |  | Good N (%) | Poor N (%) |  |  |
| Mode of Meal service | University meal service | 99 (56.7) | 582 (97.6) | 0.03 (0.01, 0.06) | 0.07 (0.02-0.22)* |
|  | Non university meal service | 76 (43.4) | 14 (2.3) | 1 | 1 |
| Monthly pocket money (ETB) | <500 | 9 (3.4) | 256 (42.9) | 0.07 (0.03, 0.14) | 0.29 (0.13, 0.64)* |
|  | >501 | 166 (94.8) | 340 (57.05) | 1 | 1 |
| Main source of meal | University meal | 40 (22.8) | 559 (93.8) | 0.01 (0.01-0.03) | 0.02 (0.01, 0.05)* |
|  | Other than university meal | 69 (39.4) | 21 (3.5) | 0.79 (0.38, 1.65) | 0.07 (0.02, 0.22) |
|  | Both | 66 (37.7) | 16 (2.6) | 1 | 1 |
| Meal outside university cafeteria | 1-2 meals a day | 57 (32.5%) | 386 (64.7) | 0.016 (0.007-0.039) | 0.20 (0.06, 0.68)* |
|  | ≥3 times a day | 57 (32.5) | 91 (15.2) | 0.06 (0.02, 0.16) | 0.55 (0.16, 1.91) |
|  | None | 6 (3.4) | 113 (18.96) | 0.006 (0.002-0.019) | 0.11 (0.02, 0.51)* |
|  | All meals | 55 (31.4) | 6 (1.00) | 1 | 1 |
